# Investigating Workplace Bullying, Intention to Quit and Depression Among Nurses in the Upper West Region of Ghana

**DOI:** 10.1101/2024.05.28.24307871

**Authors:** Emmanuel Dapilah, Andrews Adjei Druye

## Abstract

**Background:** Intention to quit among nurses is increasingly recognized as a serious predictor of voluntary turnover. Voluntary turnover on the other hand is a significant factor fueling the shortage of nurses globally which could partly be blamed on negative workplace behaviors including but not limited to workplace bullying. Even though the relationship between workplace bullying and the intention to quit has been studied extensively, little is known about these concepts among nurses in Ghana.

**Aim:** The purpose of this study was to establish the relationship between workplace bullying among nurses and their intention to quit the profession in the Upper West Region of Ghana. We also determined the relationship between workplace bullying and depression among nurses.

**Methods:** We employed a cross-sectional design with 323 nurses recruited through a multistage sampling technique. Data were collected using a structured questionnaire with a 98.5% (N=318) response rate.

**Results:** Initial descriptive statistics indicate that 64.4% (n=203) of the nurses had intentions of quitting the job while 52.1% (n=164) were depressed at various degrees based on scores on the DASS-21. Further analysis shows a positive linear relationship between WPB and intentions to quit. WPB was also correlated positively with depression among the nurses. This implies that an increased incidence of bullying at work is associated with increased intention to quit and depression among the nurses.

**Conclusions:** With over 50% of the nurses in this study having intentions of quitting the job, it would be incumbent on nurse managers and other leaders at these health facilities to reconsider the work environment, policies, and leadership to prevent actual voluntary turnover. Managers must also fashion out pragmatic strategies aimed at reducing stress and promoting the health and well-being of the nurses.

## Introduction

Nurses face daunting challenges at the workplace daily. One of these challenges at the health facilities is bullying among nurses. Workplace bullying has affected many nurses and continues to affect many more physically and psychologically. Workplace bullying poses a myriad of consequences to nurses, including but not limited to depression (1) and intention to quit (2). Also, the intention to quit is a primary determinant of voluntary turnover. Voluntary turnover, on the other hand, is fueling the global shortage of nurses amid an accelerating aging population and the burden of chronic non-communicable diseases (3).

## Workplace bullying

Workplace bullying is conceptualized as repeated behaviors perpetrated by members of an organization that are considered offensive, often escalating in intensity with a perceived intent to harm (4). It is frequently seen as a work environment well-being and security danger (5, 6). However, it remains an integral part of a more extensive complex marvel that incorporates acts of commissions or omissions in the facilities (7). While a few activities might be obvious, others will, in general, be covert, for example, refusing to intercede or retaining imperative information when activities are demonstrated and required for work to be done in a protected way (8). Several studies have found that workplace bullying is a common occurrence among nurses where it creates a harmful and fearful environment (2, 8, 9).

It has been found that employees who experience bullying at work display higher levels of burnout and turnover intentions and lower levels of job satisfaction (10). Negative behavior in the workplace can lower individuals’ psychological well-being and cause mental health problems such as anxiety, depression, lower self-esteem, and even post-traumatic stress disorder (11). Bullied employees also experience health complaints such as chronic fatigue, loss of sleep, difficulty focusing, and indigestion problems (4, 12). Additionally, bullying at work may be an antecedent of depression (13). It has also been established that workers who are victims of bullying have a higher incidence of ill health and related issues compared to those not bullied (9). Numerous factors, including the stressful nature of the act, can account for this. Initially, victims of bullying may complain of such reactions as being worried, distressed, and confused (14). With time, if the bullying behavior is not curtailed, physical and mental signs and symptoms become more evident and intense (9).

Reports of a rise in the cases of workplace bullying coupled with the increased relative risk of developing depression have ignited the claim that workplace bullying remains the most devastating psychosocial workplace exposure (15). A report from a meta-analysis by Nielsen, Magerøy (16) indicates that exposure to negative behaviors at work predisposed employees to psychiatric and physical complaints. As can be seen, the repercussions of workplace bullying are multi-faceted and traverse physical and mental disturbances, including the use of illegal substances, perpetual anger, and psychological instability (17, 18). These could lead to reduced work output, strained relationships, increased time off for treating affected staff, and poor performance.

Bullying can negatively affect the quality of concentration of employees (19), and long-term exposure may lead to a decline in the ability to stay focused at work and a loss of job satisfaction. Decreased ability to concentrate might result in medical errors and harm to patients. This is in tandem with the findings by Paice and Smith (20), who indicated that when physician trainees are bullied at work, the propensity of committing very grievous or potentially grievous medical errors remains high. Furthermore, bullying is one of the negative behaviors that has been shown to disrupt peaceful co-existence between coworkers and mar the relationships between managers and workers (21). The interpersonal relationship between nurses and patients and the relationship required for therapeutic communication is also strained, resulting in poor health outcomes and quality of care. Also, when trust is eroded in a bullying workplace, the facility bears the brunt as employees will be reluctant to highlight or report pitfalls that occur during work.

## Workplace Bullying, Intention to Quit and Voluntary Turnover Among Nurses

Intention to quit and turnover are two related yet different phenomena. The intention to quit is conceptualized as a worker’s voluntary and conscious decision to exit an organization or job, while turnover, on the other hand, is mainly described as the movement of employees across the boundaries of an organization (22). Turnover, therefore, represents workers exiting their chosen professions for other jobs. Though these concepts differ, intention to quit is the number one predictor of voluntary turnover (22, 23). In the view of Cho and Lewis (24), turnover intention is essential for two main reasons: it is a predictor of actual staff turnover and a signal that employees might not contribute to an organization at their full potential. The fact that an employee has decided to quit a job comes with much psychological strain, which tends to reduce his interest in the job and overall work output. Furthermore, the intention to quit is one of the critical indicators of staff well-being (25), which impacts the quality of patient care.

It is unclear why some employees sometimes intend to quit their current jobs. However, many factors may account for a nurse’s intention to quit, albeit inconsistent and inconclusive (26). Fochsen, Sjögren (27) found that limited opportunities, lack of professional independence, and unsatisfactory remunerations were responsible for nurses willing to quit the profession. It was also identified that men, young nurses, and nurses with muscle and joint problems were some of the predictors of nurses intending to leave the profession (28). Additionally, Griffeth, Hom (29) assert that people less satisfied with their current jobs will likely leave the profession.

When nurses have intentions to leave the job, their behavior at work and subsequent work output is affected negatively (30). Voluntary turnover is of paramount importance to facilities providing healthcare globally and is considered a serious problem that impacts hospitals, wards, and individual providers (31). Excessive turnover among nurses reduces the health facility’s ability to meet the complex and diverse needs of patients and ensure high-quality care (32). It tends to also negatively impact health outcomes for patients since it increases the loss of experienced nurses (33). Staff turnover affects organizations negatively, and as Schalk and colleagues put it, voluntary turnover is fueling the shortage of nurses across the globe and is now seen as a crisis because of its scope and resultant decline in the quality of care (34).

It is worth noting that people intending to quit represent a significant precursor to them leaving the profession. Hence, intentions to quit are used as a yardstick to quantify nurse turnover rates (35). Staff turnover leads to the loss of experienced nurses, which can compromise the safety and well-being of patients (36) and, ultimately, quality care outcomes. There are suggestions that the estimated costs related to turnover could be more than 5% of the yearly budget of most facilities (37). This places a substantial financial burden on the affected institutions, negatively influencing staff incentives and compromising the quality of care amid limited resources. Several researchers have studied the association between workplace bullying and turnover intentions and found a positive linear association between these variables (38–41).

## Relationship Between Workplace Bullying and Depression Among Nurses

Over the years, depression and anxiety disorders have been identified as common mental ailments that pose dangers to individuals and society (42). 7.8% of people from Europe are affected by a mood disorder, and 14% by anxiety disorder annually (43). Many reasons have been ascribed to the high prevalence of mental health conditions among the general population. In a survey conducted by Hansson, Chotai (44), 33% of the patients with mood disorders attributed their mental problems to circumstances at the workplace, placing issues in the work environment as the number one cause of depression among employees. The fact that work is integral to employees’ psychological health is not far-fetched, as they spend many hours daily at work.

Work or employment is a two-edged sword. While it offers remuneration, meaning, and avenues for social interactions, it can also be a source of great stress (45). Apart from work posing as a source of stress to the worker, other work-related factors and harmful workplace social acts can poison the work environment, thereby worsening the situation. Among the many work-related factors implicated in workers’ mental health problems is the workplace bullying phenomenon. In consonance with the stress theories, workplace bullying is considered a major stressor among employees, which can cause ill health and compromised well-being (46). On an individual level, workplace bullying affects all aspects of a worker’s being and might result in health-related issues such as severe headaches, depression, and anorexia (47).

According to Quine (41), nurses who became victims of workplace bullying experienced high anxiety levels, distress, and depression. Also, a study investigating workplace bullying and depression in physicians established that instances of bullying at work correlated positively with the risk of developing depression (48). Furthermore, another study conducted in Denmark concluded that there was an increased risk of depression among employees who were frequently bullied (49). Numerous studies have found a positive linear relationship between workplace bullying and depression among nurses (39, 50, 51). According to the literature, workplace bullying could lead to intention to quit and depression among nurses. The intention to quit and depression among nurses compromise their health and the quality of work output and patient care. However, there is scant research on these concepts in most developing countries, including Ghana. This study, therefore, sought to fill this knowledge gap by studying the relationship between workplace bullying and the intention to quit the job among 323 nurses working in the Upper West Region of Ghana. The study also determined the association between workplace bullying and depression among the nurses. It is essential to study nurses’ intentions to quit so that retention strategies can be drafted and implemented to reduce and/or prevent voluntary turnover.

## Methods

### Study Design

We conducted this study using a descriptive cross-sectional design, a non-experimental quantitative approach. This design was chosen because data was collected at one point (52) primarily to determine prevalence and establish associations between the study variables (53). This study was conducted among a sample of 323 Registered General Nurses and Enrolled Nurses working in five public hospitals in the Upper West Region (UWR) of Ghana. The sampling of study respondents was done through a cluster sampling technique, which is a probability method. Our choice of this technique was informed by the fact that it would have been challenging to track nurses across the region individually. As a result, we treated nurses in each district hospital as a cluster. The final sample was made up of 46.7% (n=151) Registered General Nurses (84 males and 67 females) and 53.3% (n=172) Enrolled Nurses (83 males and 89 females), which summed up to 323 nurses (167 males and 156 females).

### Data Collection Instruments

We collected the data using a structured self-administered questionnaire. A questionnaire was used to collect the data because, according to Leavy (52), questionnaires are less exorbitant and require less time and energy to administer, ensure anonymity or, to some extent, perceived privacy, and minimize response bias. This questionnaire has four sections: demographic variables, workplace bullying, depression among nurses, and intention to quit (see Appendix A).

#### Demographic variables

The demographic variables of the respondents collected in this study include gender, age, professional background, and rank/position in the profession.

#### Prevalence of workplace bullying

We adopted the Negative Acts Questionnaire-Revised (NAQ-R) developed by Einarsen, Hoel (4) to measure bullying. Total scores on bullying ranged from 22 to 110 points. Concerning current developments on the use of cutoff criteria in determining targets of workplace bullying, respondents with a cutoff score less than 33 were classified as not bullied, respondents with scores between 33 and 44 were considered occasionally bullied or targets of workplace bullying, while respondents with a score of 45 and above were considered victims of workplace bullying (54). The scale’s reliability, as tested by Cronbach’s alpha in this study, was 0.929, indicating excellent internal consistency.

#### Level of depression among nurses

The level of depression was measured with Depression Anxiety Stress Scale version 21 (DASS-21), which is a short form of DASS-42 developed by Lovibond (55) to assess the core symptoms of depression, anxiety, and stress. The DASS-21 is made up of 21 questions which are divided into three categories (Depression, Anxiety, and Stress) of seven items, each measured over the past week, and scores range from 0, “Did not apply to me at all,” to 3, “Applied to me very much, or most of the time.” The Depression sub-scale looks at hopelessness, low self-esteem, and low positive affect. The Anxiety sub-scale measures autonomic arousal, physiological hyperarousal, and the subjective feeling of fear. The Stress sub-scale items consider tension, agitation, and negative affect. For each subscale, the scores for the identified items are summed. Because DASS-21 is a short version of the original 42-item scale, the final score of each subscale (Depression, Anxiety, and Stress) was multiplied by two (x2) and then compared with the DASS Severity Ratings (see Table 1).

**Table 1.** DASS Severity Ratings.

| Severity | Depression | Anxiety | Stress |
| --- | --- | --- | --- |
| Normal | 0-9 | 0-7 | 0-14 |
| Mild | 10-13 | 8-9 | 15-18 |
| Moderate | 14-20 | 10-14 | 19-25 |
| Severe | 21-27 | 15-19 | 26-33 |
| Extremely severe | 28+ | 20+ | 34+ |

Source: Lovibond (1995).

Cronbach’s alpha for the DASS-21 and depression subscales for this study were 0.85 and 0.74, respectively.

#### Intention to quit

Finally, intention to leave (quit) was measured with a single-item, five-point scale constructed by Einarsen, Hoel (4). The respondents were required to answer the question: *“Have you considered quitting your present job over the last six months?”* based on responses that ranged from 0 *(Never)* to 4 *(Very often)*.

### Ethical Considerations

Ethical clearance was obtained from the University’s Institutional Review Board, and permission was sought from the management of the various hospitals from which the participants were recruited. Furthermore, all the participants provided written informed consent by endorsing the consent form attached to the questionnaire before answering the questions. The research assistants witnessed the signing of the consent forms. Ethical considerations stipulated by the Helsinki Declarations were also strictly adhered to.

### Data Collection Procedures

We desired to maximize the response rate amid time and budgetary constraints. Therefore, considering many factors, the in-person delivery method was adopted for this study. Leavy (52) posits that in-person surveys generally occur in group settings and have the highest response rate. Two assistants were trained to help with the in-person distribution and subsequent collection of the questionnaires, which spanned from 25^th^ February 2020 to 17^th^ April 2020. A total of 323 questionnaires were distributed across the eight study sites. However, we retrieved 318 questionnaires representing a response rate of 98.5%. Also, three (3) questionnaires from the 318 with incomplete entries representing 0.9% were rejected. Hence, the final analysis was conducted with data from 315 questionnaires.

### Data Analysis

Data analysis procedures allow us to determine the answers to the research questions or hypotheses (52). The data analysis was performed in SPSS version 29, and the alpha level was set at 0.05. Initially, frequencies were run for the data, and inspection was done to identify missing values and errors. Some ordinal variables were transformed into dichotomous variables to allow for better result presentation, discussion, and communication of findings. For example, the variable ‘age’ collected as a continuous variable was transformed into a categorical variable based on the following groups: 20-29, 30-39, and 40+ years. Current professional ranks were grouped into two: junior nurses and senior nurses. All items in the questionnaire measuring workplace bullying were transformed into a single variable to facilitate data analysis. As such, the 22 items in the NAQ-R were coded and transformed into one main variable, which measured exposure to bullying. The first part of the NAQ-R has 22 items, and each item was measured on a 5-point Likert scale (1-never, 2-now and then, 3-monthly, 4-weekly, and 5-daily). The total score on the NAQ-R was, therefore, 110 (5*22=110). The total score obtained by each respondent was further categorized into three groups according to cutoff points by Notelaers and Einarsen (2013) as follows: less than 33, not bullied; 33-44, occasionally bullied or targets of bullying; and 45+, victims of bullying.

Similarly, seven (7) items on the DASS-21 measured depression. Each item was measured on a 4-point Likert scale from 0 to 3. So, the total score for depression as measured by the DASS-21 was 21 (3*7=21). However, since the DASS-21 is an abridged version of the original DASS-42, whatever score was obtained from DASS-21 was multiplied by 2 to obtain the actual depression score. Based on the actual scores, the level of depression among the respondents was categorized according to the DASS severity rating by Lovibond (55) as follows: 0-9=normal, 10-13=mild depression, 14-20=moderate depression, 21-27=severe depression, and 28+=extremely severe depression.

## Results

### Demographic Variables of Respondents

The initial analysis shows that 52.1% (n=164) were males, while 47.9% (n=151) were females. We observed that about 47.9% of the nurses were between the ages of 20 to 29 years. This was followed by about 39.4% for those between 30 and 39 years old. The least were those aged 40 years and above, representing only 12.7%. Regarding the professional background of the nurses, it was observed that 53% of them were Enrolled Nurses, while the remaining proportion (47%) were Registered General Nurses (see Table 2 for details).

**Table 2.**
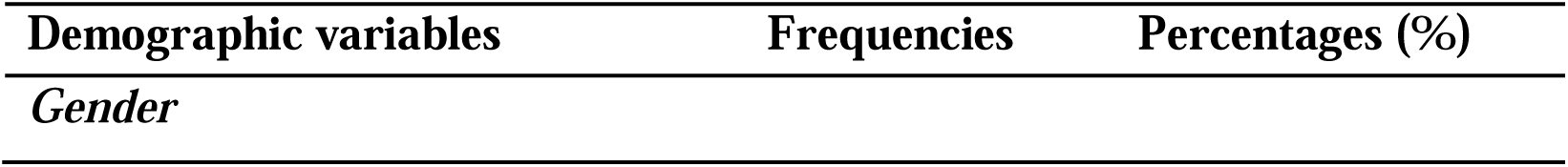
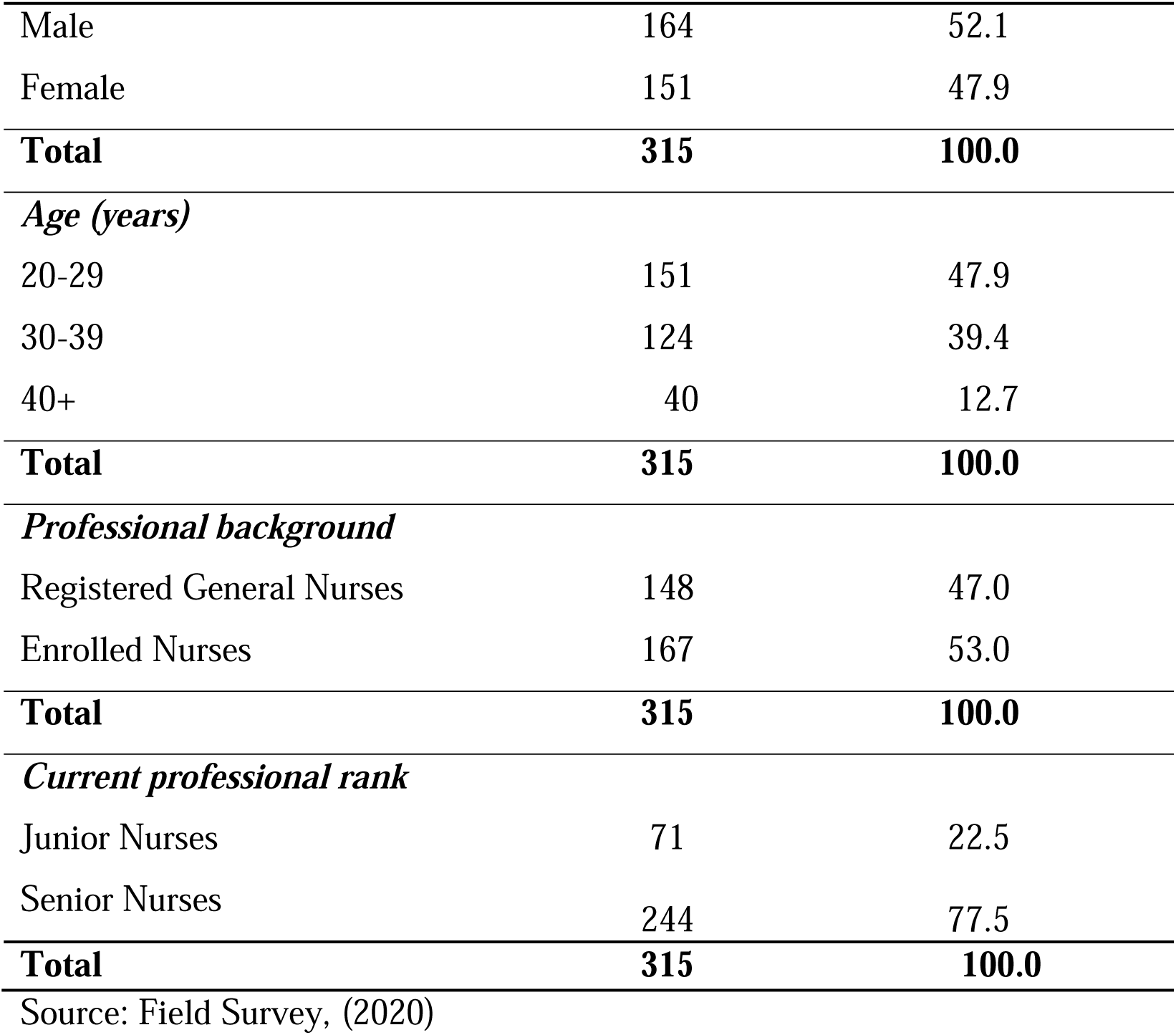
Demographic Characteristics of Respondents.

| Demographic variables | Frequencies | Percentages (%) |
| --- | --- | --- |
| Gender |  |  |
| Male | 164 | 52.1 |
| Female | 151 | 47.9 |
| **Total** | **315** | **100.0** |
| *Age (years)* |  |  |
| 20-29 | 151 | 47.9 |
| 30-39 | 124 | 39.4 |
| 40+ | 40 | 12.7 |
| **Total** | **315** | **100.0** |
| *Professional background* |  |  |
| Registered General Nurses | 148 | 47.0 |
| Enrolled Nurses | 167 | 53.0 |
| **Total** | **315** | **100.0** |
| *Current professional rank* |  |  |
| Junior Nurses | 71 | 22.5 |
| Senior Nurses | 244 | 77.5 |
| **Total** | **315** | **100.0** |

Source: Field Survey, (2020)

### Intention of Nurses to Quit the Profession

Regarding nurses’ intention to quit, 64.4% (n=203) have indicated that they have considered leaving the nursing profession within the past six months, while 35.6% (n=112) said they have not contemplated leaving the profession (see Table 3).

**Table 3.** Intention of Nurses to Quit the Profession.

|  |  | Frequency | Percent |
| --- | --- | --- | --- |
| Valid | Never | 112 | 35.6 |
|  | Rarely | 63 | 20.0 |
|  | Sometimes | 92 | 29.2 |
|  | Quite often | 32 | 10.2 |
|  | Very often | 16 | 5.1 |
|  | **Total** | **315** | **100.0** |

Source: Field Survey (2020).

### Prevalence of Depression Among Nurses

Regarding depression, 52.1% (n=164) of the nurses over the study period have experienced signs and symptoms of depression at various levels based on scores on the DASS-21 (see Table 4).

**Table 4.**
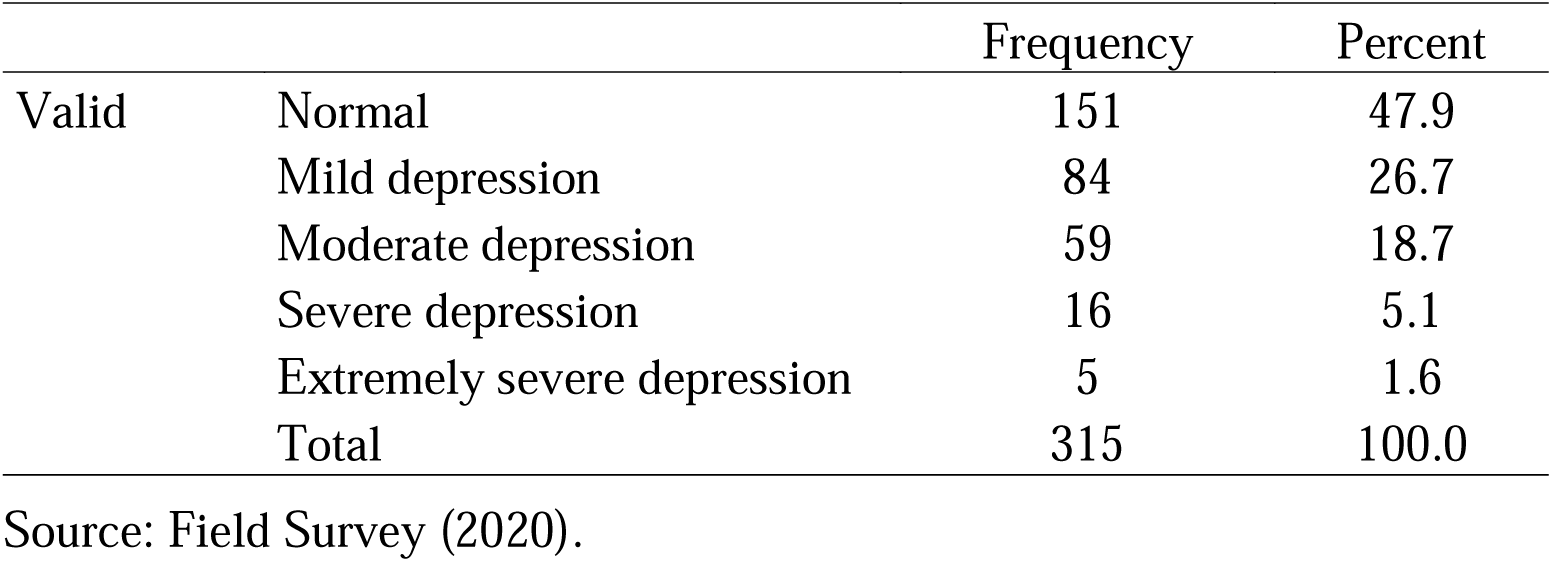
Prevalence of Depression among Nurses Based on DASS-21.

### Relationship Between Workplace Bullying and the Intention to Quit

The relationship between workplace bullying and the intention of a nurse to quit his/her job was also assessed, and the results are displayed in Table 5.

**Table 5.** Correlations of Workplace Bullying and Intentions to Quit.

Correlations of Workplace Bullying and Intentions to Quit
|  |  | Intentions of Respondents to<br>Quit Current Job |
| --- | --- | --- |
| Total perceived<br>bullying | Pearson Correlation | 0.487** |
|  | Sig. (2-tailed) | 0.000 |
|  | **N** | **315** |

Source: Field Survey (2020)
\*\*Significant at $p < 0.001$

The relationship between perceived bullying at work (as measured by the NAQ-R) and nurses’ intention to quit the profession (measured by a single item on a 5-point Likert scale) was investigated using the Pearson product-moment correlation coefficient. Initial investigations were carried out to rule out violations of the assumptions of normality, linearity, and homoscedasticity. The results show a strong, positive correlation between the two variables (r=0.487; N=315, *p*<0.001), with high levels of perceived workplace bullying associated with increasing levels of nurses’ intentions to quit their jobs. This means that as the perceived level of workplace bullying increases, the level of the intention of a nurse to quit his/her job increases and vice versa.

### Relationship Between Intention to Quit and Depression Among Nurses

We assessed the relationship between the intention of a nurse to quit his/her job and depression using Pearson correlation (see Table 6 for details).

**Table 6.** Relationship Between Intention to Quit and Depression Among Nurses.

|  |  | Actual depression score |
| --- | --- | --- |
| Intention to Quit | Pearson Correlation | 0.363** |
|  | Sig. (2-tailed) | 0.000 |
|  | N | 315 |

Source: Field Survey (2020)
\*\*Significant at $p<0.001$

The results indicate a strong, positive correlation between the two variables (r=0.363; N=315, *p*<0.001), with high levels of depression associated with increasing levels of the intentions of nurses to quit their jobs. This means that as the perceived level of depression increases, the level of the intention of a nurse to quit his/her job increases and vice versa.

### Relationship Between Workplace Bullying and Depression Among Nurses

We also determined the relationship between workplace bullying and depression among the respondents. Again, the relationship between perceived bullying at work (as measured by the NAQ-R) and depression among nurses (as measured by the DASS-21) was investigated using the Pearson product-moment correlation coefficient. Initial investigations were carried out to rule out violations of the assumptions of normality, linearity, and homoscedasticity, and the results are presented in Table 7.

**Table 7.** Relationship Between Workplace Bullying and Depression.

|  |  | Actual depression score |
| --- | --- | --- |
| Total perceived bullying | Pearson Correlation | 0.559** |
|  | Sig. (2-tailed) | 0.000 |
|  | N | 315 |
| Source: Field Survey (2020) |  | **Significant at p<0.001 |

From the analysis, there was a strong, positive correlation between the two variables (r=0.559; N=315; *p*<0.001), with high levels of perceived workplace bullying being associated with increasing levels of depression among nurses. Thus, the higher the perceived level of workplace bullying, the higher the level of depression among the nurses and vice versa.

To find out if workplace bullying and depression were significant independent predictors of nurses’ intention to quit their jobs, we conducted multiple linear regression analyses. All the assumptions of the general linear models were not violated. A unit increase in workplace bullying leads to a 0.036 unit increase in the intention to quit while controlling for depression, age, professional background, gender, and rank of the nurse (B=0.036, 95% CI 0.025; 0.046). We can be 95% confident that the true population parameter lies between 0.025 and 0.046. Also, a unit increase in depression leads to a 0.026 unit increase in the intention to quit while controlling for workplace bullying, age, professional background, gender, and rank of the nurse (B=0.026, 95% CI 0.005; 0.048). Hence, we can be 95% confident that the true population parameter lies between 0.005 and 0.048. The adjusted R^2^ for the regression model is 0.244. This indicates that 24.4% of the variance in the nurses’ intention to quit is accounted for by workplace bullying and depression. (see Table 8 for details).

**Table 8.**
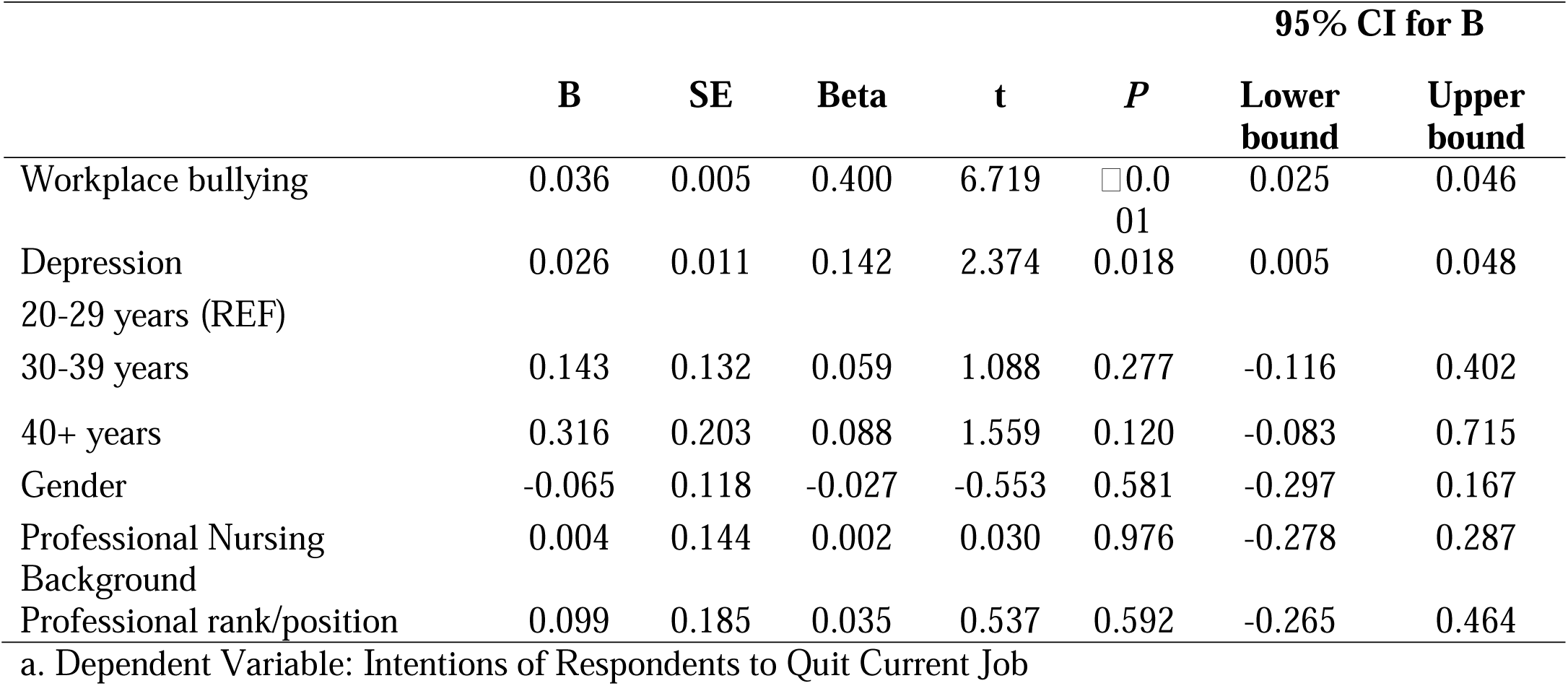
Summary of Multiple Linear Regression.

## Discussion

More than 60% of the nurses in this study have stated that they have contemplated leaving the profession within the past six months. This figure is enormous because most of the facilities in Ghana have a shortfall in the number of nurses. It was necessary to ascertain whether workplace bullying was related to the nurses’ intentions to quit their jobs. The Pearson correlation coefficient indicated that workplace bullying was positively related to nurses’ intention to quit the profession. This means increased workplace bullying prevalence could increase nurses’ intention to leave their profession. Several studies have also established a positive association between workplace bullying and nurses’ intentions to leave the profession (38–40, 56, 57).

Intention to quit could result in actual staff turnover. Staff turnover leads to the loss of qualified nurses, declining patient safety (36), and high operational expenses (58). Nurses who are victims of workplace bullying have increased chances of falling sick and experiencing other health-related issues than workers who are not bullied (9). It is also documented that employees who are victims of workplace bullying may become distressed, worried, and confused (14). Because workplace bullying is prevalent among nurses, it stands to reason that bullying will pose health and safety risks to them, which may ultimately trigger their intentions to leave the profession for other jobs that are less stressful and risky. This might help explain the positive association between workplace bullying and the nurses’ intentions to quit the profession.

In this study, over 50% of the nurses have experienced various degrees of depression according to measures on the DASS-21. However, it is important to note that DASS-21 is a quantitative measure of distress (depression, anxiety, and stress) and therefore not a definite measure of clinical diagnosis (55). What this means is that the assessment scores obtained from the DASS-21 can be used to identify individuals experiencing considerable symptoms who might be at high risk of developing clinical depression. There is, therefore, a need to refer such people for further psychological evaluation.

Further investigations were conducted on the relationship between workplace bullying and depression among nurses who took part in the study. It was determined from the Pearson correlation coefficient that there was a strong positive linear relationship between workplace bullying and depression among the nurses. This is an indication that any increase in workplace bullying prevalence subsequently would be associated with an increase in the nurses’ depression states and vice versa. This finding is consistent with earlier studies (2, 39, 51, 59), which found a positive linear relationship between workplace bullying and depression.

Work offers livelihood for employees and, at the same time, can be seen as a source of great stress. According to Hoobler, Rospenda (46), workplace bullying has been recognized as a primary source of distress that is associated with subsequent ill-health and decreased well-being. As such, anything that negatively affects work or the working environment will inadvertently increase employees’ stress levels and result in compromised well-being. This might account for the positive linear relationship between workplace bullying and depression. However, the findings in this study contradict those of Doe (60) who found no significant association between workplace bullying and depression among a sample of university workers in Ghana. The reasons for the difference in findings are not readily known. Hence, further research is required to help elucidate these findings.

The multiple linear regression results have indicated that workplace bullying (B=0.036, 95% CI=0.025; 0.046) and depression (B=0.026, 95% CI=0.005; 0.048) are independent predictors of nurses’ intention to quit their jobs even after adjusting for the nurses’ age, gender, professional background, and position. The whole regression model accounted for 24.4% of the variance in the intention to quit. This means that although workplace bullying and depression are significant factors in the decision of nurses to quit their profession, there are other reasons not captured by this study. We can speculate that low salaries and remunerations, low prestige, discrimination, and negative public perceptions about the nurse could be accountable for nurses intending to quit their jobs in Ghana. These are similar to the reasons identified by Fochsen, Sjögren (27), who found a lack of professional opportunities, restricted professional autonomy, and unsatisfactory salary as precursors to the decision to quit. The intention of workers to quit their professions has been found to predict actual staff turnover strongly, and this indicator has been used widely in nurse turnover studies (35). However, with the high unemployment levels among newly qualified nurses and the unavailability of alternative job opportunities for nurses in Ghana, we are uncertain whether the intention to quit would result in actual nurse turnover. Therefore, longitudinal studies need to be conducted to determine if the intention to quit among nurses will lead to staff turnover.

## Conclusions

Workplace bullying, intention to quit, and depression are prevalent among nurses in this study area. There is evidence that a substantial number of nurses in this study are considering quitting their current job as nurses, and most of them are depressed. The intention to quit among the nurses was found to be predicted by bullying at the facilities and depression among the nurses. Once it is established that workplace bullying and depression are independent predictors of the nurses’ intention to quit, measures must be instituted to curb the occurrence of bullying at work and depression among the nurses. When there is a reduction in the incidence of bullying at work, the chances that workers will desire to quit their jobs or become depressed might be minimized. This will create a cordial and productive work environment, ultimately leading to quality care outcomes and a healthy hospital workforce.

## Recommendation for further research

Considering the number of nurses in this study who have the intention to quit their jobs amidst the high unemployment among newly qualified nurses and the lack of alternate job opportunities for nurses in Ghana, we recommend that researchers carry out longitudinal studies to establish whether the intention to quit one’s job will result in actual staff turnover. There is also a need for studies that will comprehensively study the factors responsible for nurses intending to leave the profession since the current study accounted for only 24% of the variance in the intention to quit.

## Data Availability

All relevant data are within the manuscript and its Supporting Information files.

https://rochester.app.box.com/folder/0

## Key Points for Policy, Practice, and/or Research

1. A greater number of nurses are considering quitting their jobs. This unfortunate situation is partly attributed to workplace bullying and depression. Therefore, measures must be taken to curb workplace bullying and prevent nurses from leaving the profession.
2. This study has highlighted the need to regularly screen all nurses in our facilities since most of them are exhibiting signs and symptoms of depression. The mental health of nurses must be taken seriously and must be incorporated into the annual health screening exercise of hospitals.
3. Since several nurses are willing to quit the profession without alternative job opportunities in Ghana, longitudinal studies are needed to determine whether the intention to quit will result in actual staff turnover.

## Author Biographies

Emmanuel Dapilah is a highly motivated Nurse and PhD student at the University of Rochester. He is interested in the development of high standards of nursing care and patient safety.

Andrews Adjei Druye is a great mentor, a nurse researcher, and a senior lecturer at the School of Nursing and Midwifery, Department of Adult Health at the University of Cape Coast.

## Acknowledgement of financial support

The corresponding author received financial support from the Samuel and Emelia Brew-Butler-SGS/GRASAG UCC Research Fund 2020 while preparing the thesis submitted to the University of Cape Coast in partial fulfillment of the requirements for the award of Master of Nursing (MN).

## Conflict of interest statement

We declare that there are no conflicts of interest in this research report.

## Competing interests

The authors have no competing interests to declare.

## Authors contributions

ED conceived, designed, and drafted the study. AAD supervised data collection and analysis and critically reviewed and approved the final manuscript.

## Notes

### Competing Interest Statement

The authors have declared no competing interest.

### Clinical Protocols

https://rochester.app.box.com/folder/0

### Funding Statement

The author(s) received no specific funding for this work.

### Author Declarations

Ethical clearance was obtained from the University of Cape Coast Institutional Review Board (UCCIRB/CHAS/2019/209), and permission was sought from the management of the various hospitals from which the participants were recruited. Furthermore, all the participants provided written informed consent by endorsing the consent form attached to the questionnaire before answering the questions. The research assistants witnessed the signing of the consent forms. Ethical considerations stipulated by the Helsinki Declarations were also strictly adhered to.

